# Alcohol intake and hypertensive disorders of pregnancy: a negative control analysis in the ALSPAC cohort

**DOI:** 10.1101/2021.12.02.21267176

**Authors:** Florence Z Martin, Abigail Fraser, Luisa Zuccolo

## Abstract

**Introduction:** Alcohol intake increases blood pressure, yet estimates of associations between maternal intake and hypertensive disorders of pregnancy (HDP) are sparse and range from null to a protective effect. Here we estimated the association of maternal drinking during pregnancy with preeclampsia and gestational hypertension (separately and jointly, as HDP). We used partner’s alcohol intake as a negative control exposure, beverage type-specific models, and a range of sensitivity analyses to strengthen causal inference and reduce the influence of bias.

**Methods:** We performed a prospective cohort study using data on self-reported alcohol intake in the UK Avon Longitudinal Study of Parents And Children (ALSPAC) and HDP ascertained from obstetric notes. Multivariable multinomial regression models were adjusted for confounders and mutually adjusted for partner’s or maternal alcohol intake in the negative control analysis. We also performed a beverage type analysis of the effect of beer and wine separately on HDP risk, due to different social patterning associated with different drinks. Sensitivity analyses assessed the robustness of results to assumptions of no recall bias, no residual confounding, and no selection bias.

**Results:** Of the 8,999 women eligible for inclusion, 1,490 developed HDP (17%). Both maternal and partner’s drinking were associated with decreased HDP odds (mutually adjusted odds ratio 0.86, 95% confidence interval 0.77 to 0.96, P-value=0.008 and 0.82, 0.70 to 0.97, P=0.018, respectively). We demonstrate the validity of the negative control analyses using the same approach for smoking as the exposure. This confirmed an inverse association for maternal but not partner’s smoking, as expected. Estimates were more extreme for increasing levels of wine intake compared to increasing levels of beer. Multiple sensitivity analyses did not alter our conclusions.

**Conclusion:** We observed an inverse relationship between alcohol intake during pregnancy and risk of HDP for both maternal and, more surprisingly, partner’s drinking. We speculate that this is more likely to be due to common environmental exposures shared between pregnant women and their partners, rather than a true causal effect. This warrants further investigation using different study designs, including Mendelian randomisation.

## Introduction

Hypertensive disorders of pregnancy (HDP) is an umbrella term for gestational hypertension and preeclampsia, both characterized by *de novo* hypertension arising during pregnancy, with concurrent proteinuria in preeclampsia (1). There are several known risk factors for the development of HDP, screened for at the antenatal booking appointment, including older maternal age, obesity, history of HDP, and diabetes (2). While alcohol intake is known to increase blood pressure (3-6), previous studies have produced inconsistent results regarding the risk of HDP when comparing women consuming alcohol in pregnancy to those abstaining (7-10).

In the absence of randomised controlled trials or natural experiments investigating the role of alcohol on HDP, relevant evidence comes entirely from observational studies. Residual confounding by factors such as socio-economic position and smoking is a concern, since smoking and drinking alcohol are correlated (11) and socially patterned, and smoking during pregnancy is associated with a lower risk of developing preeclampsia (12). Therefore, failure to adequately account for smoking in analyses of the association between prenatal alcohol and preeclampsia and HDP could lead to biased estimates in the same direction as the smoking-HDP effect.

A recent (not currently peer-reviewed) systematic review showed some evidence of an inverse association between alcohol use in pregnancy and preeclampsia, especially when examining prospective studies (pooled odds ratio (OR) 0.64, 95% confidence interval (CI) 0.54 to 0.76) (13). The evidence pointing to an inverse association is paradoxical given the blood pressure-elevating effect of alcohol intake outside of pregnancy.

Negative control designs can be utilised in observational epidemiological studies to elucidate whether an association is likely to be causal or whether it is a result of unmeasured or residual confounding (14). For studies examining exposures during pregnancy, partner behaviours can be used as the negative control exposure for maternal outcomes. This is based on the assumption that partner’s alcohol intake should not cause maternal HDP. If an association is observed, it suggests a common confounding structure by shared environment.

In this study, we aimed to quantify the association between alcohol intake during pregnancy and HDP in a large population-based cohort – the Avon Longitudinal Study of Parents And Children (ALSPAC). We employed a negative control exposure design, using partners’ alcohol intake during pregnancy, to detect the presence of confounding and disentangle association from causation. We also performed a beverage type analysis of the effect of beer and wine separately on HDP risk, due to different social patterning associated with different drinks, and a range of sensitivity analyses to increase confidence in our findings.

## Methods

### Participants and recruitment

We used information from the ALSPAC cohort to define the study population in this analysis. ALSPAC is a UK-based cohort of 15,454 women recruited in the early nineties from the Southwest of England and followed up pre- and postnatally via self-report questionnaires and in-person clinics. Previous publications have described the maternal cohort in full (15) and several online resources are available to browse ALSPAC data (http://www.bristol.ac.uk/alspac/researchers/our-data/) (16). We included mothers with self-report questionnaire data on alcohol intake during pregnancy and other covariates deemed to be potential confounders, as well as obstetric data abstracted from medical records (*n*=8,999) (Figure 1).

**Figure 1.**
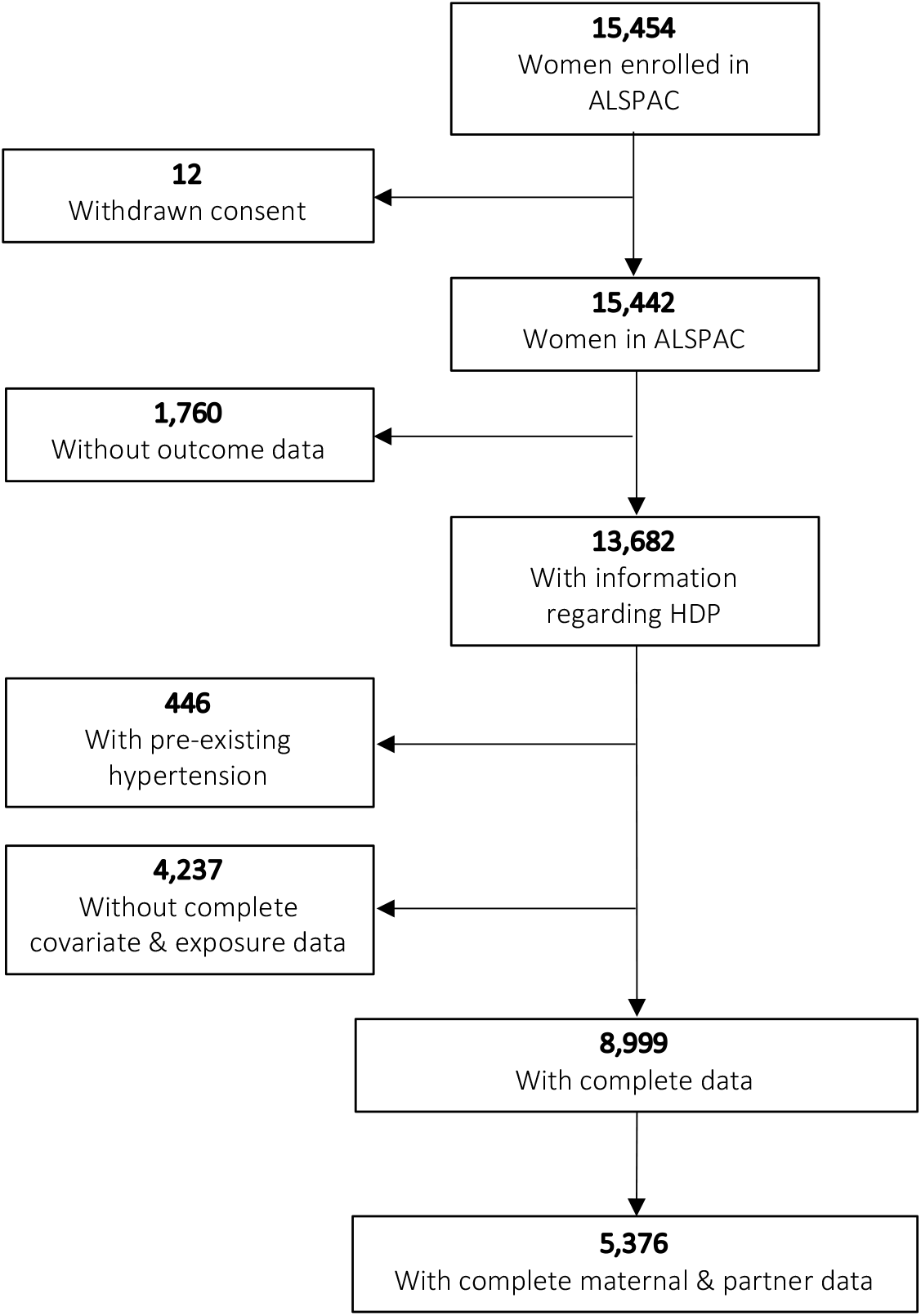
Flow of participants through the study.

Ethical approval for this study was secured from the ALSPAC Ethics and Law Committee and local Research Ethics Committee (North Somerset and South Bristol). Participants gave consent for their obstetric data to be abstracted and answers to self-report questionnaires to be used in subsequent research; individuals have the right to withdraw from the ALSPAC cohort at any time during follow-up.

## Measures

### Alcohol consumption in pregnancy

The exposure in this study, alcohol intake during pregnancy, was measured using multiple questionnaires sent prenatally and in the immediate postpartum period. At around 18 weeks’ gestation, participants were asked how often they drank alcohol: (i) in the first three months of pregnancy and (ii) since the baby first moved. These questions were categorized as: none, <1 drink per week, 1+ glasses per week, 1–2 glasses per day, 3–9 glasses per day, and 10+ glasses per day. They were also asked about how much of each type of drink (beer, wine, spirits or other) they drank on a typical day, having been advised that a glass was the equivalent of a half pint (beer), a wineglass (wine), or a pub measure (spirit). The questionnaire that was sent at the same time to partners asked the same questions regarding alcohol intake. After birth, mothers and partners were asked about average alcohol intake in the final two months of pregnancy, using the categories listed above. Using the answers given in these questionnaires, the maximum amount of alcohol that each participant reported to drink at any time during pregnancy was used to categorise women: none, low-to-moderate (1–6 drinks per week), and heavy (≥7 drinks per week).

At 18 weeks’ gestation, both mothers and partners were asked how many days in the last month they had consumed the equivalent of two pints of beer or more. Although this does not perfectly align with other definitions of binge drinking (17) including the National Institute for Alcohol Abuse and Alcoholism’s definition (≥4 drinks in two hours) (18), it provided an appropriate additional category for sensitivity analyses to separate those in the “heavy” drinking category who were not bingeing with those who were drinking multiple alcoholic beverages in one day.

Given the specific questions asked at 18 weeks’ gestation pertaining to the intake of different types of alcoholic beverages at the time of the questionnaire being filled out, we derived two variables for beer and wine intake during pregnancy. In other words, the beer drinker group consisted of those who had not reported wine consumption and vice versa for wine. We used the same categorisation of amounts drank as the primary analysis (none, low-to-moderate, and heavy) for each beverage type. Reporting of spirits/other alcohol intake and bingeing was then compared in beer and wine groups to better understand overall drinking patterns in these two groups.

### Hypertensive disorders of pregnancy

For women who gave informed consent to have their obstetric data abstracted, all recorded measurement of both systolic and diastolic blood pressure, as well as events of proteinuria, as previously described in detail (19). Women were then categorized as: normotensive, gestational hypertension, or preeclampsia, as per the 1988 International Society for the Study of Hypertension in Pregnancy criteria. (1). As shown in Figure 1, women with existing hypertension were excluded.

### Other variables

Covariates for this analysis were defined *a priori* using evidence from the literature to support a potential relationship with both the exposure and the outcome: maternal age at delivery, maternal ethnicity, maternal body mass index (BMI), smoking status (before and during pregnancy), maternal socioeconomic position (SEP), marital status, and parity. Women reported their age, ethnicity, height and weight (used to calculate pre-pregnancy BMI), smoking habits, educational attainment (proxy for SEP), marital status, and parity on self-completed questionnaires sent out during pregnancy.

Three questionnaires asked participants about their smoking habits at different times during pregnancy: at 18 weeks’ gestation women were asked about smoking early in pregnancy and current smoking, at 32 weeks’ gestation current smoking habits were described, and at 8 week’s postpartum participants reported their smoking habits in the last two months of pregnancy. Two smoking variables were generated: a binary variable for any or no smoking during pregnancy and a categorical variable for average number of cigarettes smoked per day during pregnancy.

All the variables described above were also measured via self-report questionnaire for the partners of participant’s, which were abstracted for adjustment of the negative control analysis. Partner’s smoking status was measured across several variables in two prenatal questionnaires, which were collated to create a binary variable of any or no smoking during their partner’s pregnancy.

## Statistical analysis

Women’s characteristics were described by levels of alcohol intake in pregnancy using means (standard deviations) for continuous variables and percentages for binary variables. There was no evidence of an association between HDP (outcome) and study attrition and given that all covariates had <15% missing data. Thus, we deemed that multiple imputation would not increase the study efficiency in this case and the use of a complete case analysis (CCA) was the most appropriate approach (20, 21) (supplementary tables 1 and 2).

For the primary analysis, we used multivariable logistic regression to estimate the odds ratio of HDP by increasing categories of alcohol intake (none, low-to-moderate, and heavy drinking). Due to the three-level exposure variable, likelihood-ratio tests were used to test for dose-response, comparing alcohol use as a single 3-level (continuous) variable (model A) or including alcohol as two dummy variables (model B). We used multivariable multinomial logistic regression models to estimate the relative risk ratio of developing gestational hypertension and preeclampsia compared with normotensive, using the outcome over three categories.

The primary analysis was then repeated using partners’ alcohol intake as the exposure. The comparison of maternal and partner’s association with HDP rested on the assumption that mothers and partners share environmental and behavioural factors affecting or correlating with their alcohol drinking which also affect maternal HDP risk, but only maternal alcohol use could physiologically affect HDP risk. Both adjusted and mutually adjusted models were fitted, with the latter additionally adjusting for partner’s or mother’s alcohol intake in turn to account for the potential bias from assortative mating (22). We additionally report the association of maternal and partner’s smoking during pregnancy with risk of HDP, with similar mutual adjustments. Smoking during pregnancy was used as an supplementary exposure in the negative control model to check our prior assumption that a maternal exposure with evidence of an association with HDP, such as maternal smoking, should indeed be associated with HDP but that partner exposure would not.

To further evaluate the role of residual confounding by SEP or associated factors, we compared estimates of the association of HDP risk with wine and beer drinking separately. This was done under the assumption that intake of these two beverages follow different SEP patterning, as previously demonstrated in this cohort (23). It follows therefore that consistent results would strengthen a causal interpretation, whereas discordant results could point to confounding biasing the findings.

We conducted sensitivity analyses to assess to what extent estimates obtained from the primary analysis were robust to sources of bias including:: (i) excluding those women who responded to alcohol-related questions after 20 weeks’ gestation to limit recall bias (HDP status influencing reporting of the exposure), (ii) using a categorical smoking covariate in the model (as opposed to binary) to better account for residual confounding by smoking, and (iii) excluding women who abstained from alcohol prior to pregnancy to limit the potential impact of existing ill-health.

## Results

### Study sample

After exclusions, 8,999 women (58% of the whole sample) were eligible for inclusion in this study (Figure 1), of whom 1,490 developed HDP (17% of the eligible sample). Table 1 shows the characteristics of included participants, by amounts of alcohol intake during pregnancy. Those who reported low-to-moderate drinking were older, more highly educated, more likely to be white and had a lower BMI, compared to those who reported no alcohol intake during pregnancy. Compared to non-drinkers, heavy drinkers were also more likely to be older, white, and more highly educated; heavy drinkers were also more likely to smoke both before and during pregnancy, had more children and were less likely to be married (Table 1).

**Table 1.** Participant characteristics by categories of alcohol intake during pregnancy (*n*=8,999)

|  | No alcohol in pregnancy | Low-to-moderate alcohol intake in pregnancy | Heavy alcohol intake in pregnancy |
| --- | --- | --- | --- |
|  | 2,415 | 4,696 | 1,888 |
| **Age at delivery** |  |  |  |
| Mean, years (SD<sup>a</sup>) | 27.7 (4.7) | 28.9 (4.6) | 28.7 (4.9) |
| **BMI<sup>b</sup> (12 weeks' gestation)** |  |  |  |
| Mean, kg/m<sup>2</sup> (SD<sup>a</sup>) | 23.0 (4.0) | 22.7 (3.6) | 23.1 (3.6) |
| **Smoking** |  |  |  |
| Any pre-pregnancy, *n* (%) | 705 (29.2) | 1,299 (27.7) | 874 (46.3) |
| Any during pregnancy, *n* (%) | 552 (22.9) | 991 (21.1) | 753 (39.9) |
| **Parity (18 weeks' gestation)** |  |  |  |
| Multiparous, *n* (%) | 1,280 (53.0) | 2,589 (55.1) | 1,124 (59.5) |
| **Ethnicity (32 weeks' gestation)** |  |  |  |
| Non-white, *n* (%) | 61 (2.5) | 80 (1.7) | 24 (1.3) |
| **Educational attainment (32 weeks' gestation)** |  |  |  |
| University degree, *n* (%) | 232 (9.6) | 792 (16.9) | 221 (11.7) |
| **Marital status** |  |  |  |
| Married, *n* (%) | 1,904 (78.8) | 3,792 (80.8) | 1,303 (69.0) |

<sup>a</sup>Standard deviation
<sup>b</sup> Body mass index

### Maternal alcohol intake and HDP

Figure 2 shows the association of maternal alcohol intake during pregnancy with HDP in women with complete data (*n*=8,999). The likelihood ratio test comparing model A with model B showed that the more parsimonious model A (alcohol as a 3-level continuous variable) provided as good a fit to the data as model B (alcohol as two dummy variables) (P-value=0.87), thus no evidence of a non-linear association. A one category increase in alcohol intake was associated with lower odds of developing HDP (adjusted OR 0.85, 95% CI 0.78 to 0.92, P<0.001). Similarly, the adjusted relative risk ratio for the multinomial logistic regression was 0.86 (0.79 to 0.94, P=0.001) for gestational hypertension and 0.74 (0.59 to 0.92, P=0.007) for preeclampsia.

**Figure 2.**
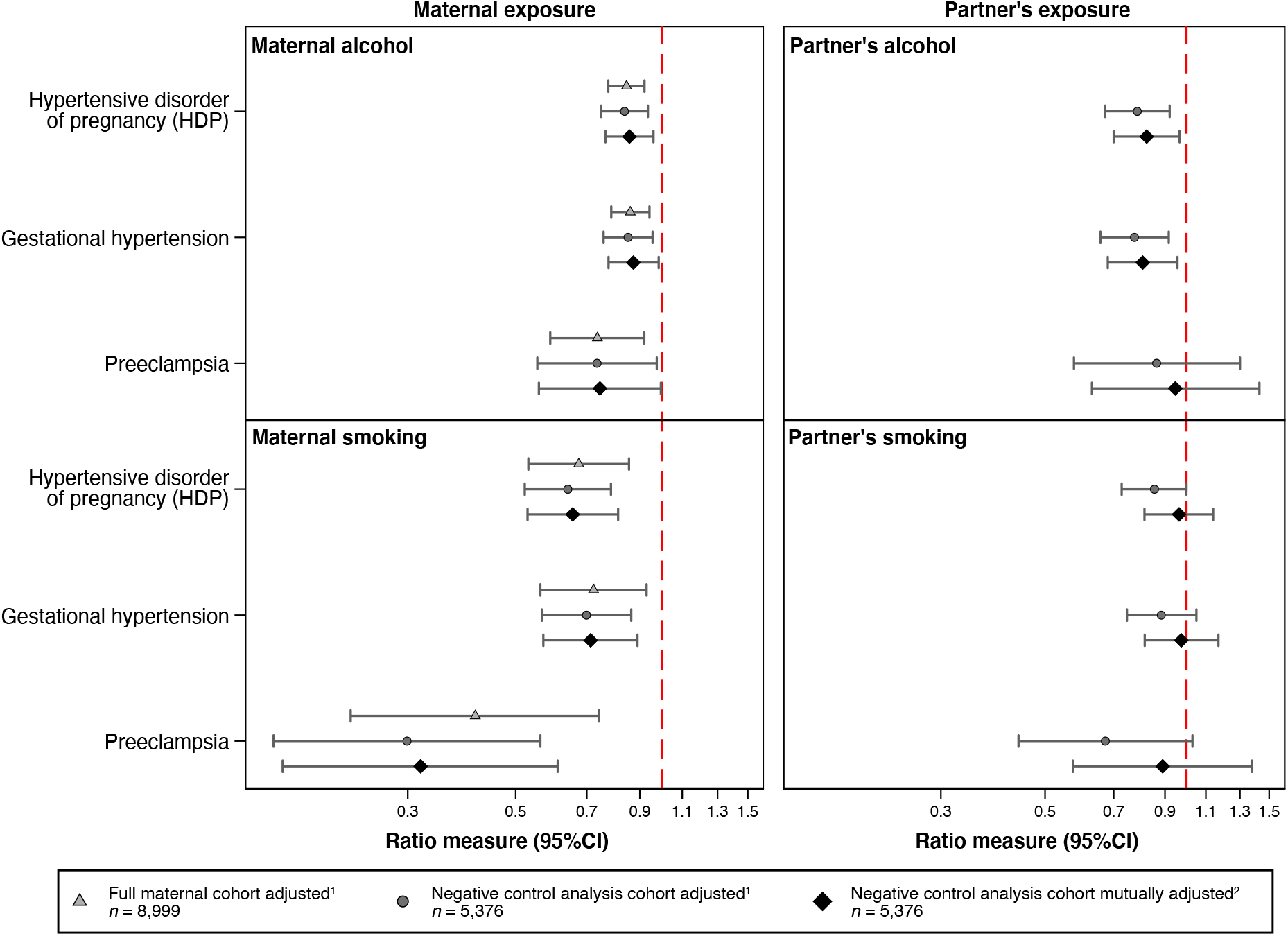
Primary and negative control analysis showing associations between maternal alcohol intake and smoking during pregnancy, as well as partner’s alcohol use and smoking during pregnancy, and maternal HDP, gestational hypertension and preeclampsia. Association between alcohol and smoking during pregnancy in mothers and partners. One category increase in maternal alcohol intake (non-drinker, low-to-moderate or heavy drinker), is associated with a decreased odds of developing HDP in both the full cohort and the negative control cohort, both adjusted and mutually adjusted models (mutually adjusted odds ratio 0.86, 95% confidence interval 0.77 to 0.96). Similarly, partner’s drinking (in the same increasing levels as described for maternal alcohol intake) is associated with a decreased odds of HDP in the adjusted and mutually adjusted model. Any maternal smoking during pregnancy (smoker or non-smoker) shows a strong negative association with HDP in all cohorts and models, as compared with no smoking; partner’s smoking during pregnancy however is not associated with maternal HDP risk when mutually adjusting for maternal smoking. ^1^ Adjusted for age, BMI, smoking (in the alcohol model), alcohol (in the smoking model), parity, ethnicity, educational attainment, and marital status (maternal or partner covariates depending on the exposure model). ^2^ Mutually adjusted for all covariates in the adjusted models plus mother/partner alcohol intake/smoking (depending on the exposure model).

When restricting to the sample of pregnancies with complete data on both mothers and partners (*n*=5,376) (Figure 1), which we refer to as the negative control cohort, we obtained similar results that persisted after mutual adjustment (mutually adjusted OR 0.86, 0.77 to 0.96, P=0.008).

Heavy drinkers were split into heavy non-binge and heavy binge drinking to ascertain whether the protective effect may be driven by those drinking “little and often”. Both binge and non-binge drinking were inversely associated with HDP, and confidence intervals overlapped between drinking categories (supplementary tables 5 and 6).

### Negative control analysis using partner’s alcohol intake

In adjusted analyses, there was evidence that partner’s drinking was associated with maternal HDP risk even after mutual adjustment for maternal drinking (mutually adjusted OR 0.82, 95% CI 0.70 to 0.97, P-value=0.018, Figure 2).

An inverse association was observed with gestational hypertension, however there was little evidence of association of partner’s drinking with preeclampsia (0.95, 0.63 to 1.43, P=0.79, Figure 2). The number of partners who were non-drinkers was lower than the number of mothers, resulting in a smaller number of preeclamptic pregnancies in that exposure category (Figure 2, supplementary table 7).

### Negative control analysis using smoking during pregnancy

As shown in Figure 2, we found evidence that maternal smoking during pregnancy was strongly associated with lower HDP risk in both the full and negative control cohort, with results almost unchanged after adjusting for partner’s smoking (mutually adjusted OR 0.66, 95% CI 0.53 to 0.81, P-value<0.001). We found similar results for gestational hypertension (0.71, 0.57 to 0.89, P=0.003), and a stronger association for preeclampsia (0.32, 0.17 to 0.61, P=0.001). On the other hand, adjustment for maternal smoking impacted the estimates for partner’s smoking. Based on mutually adjusted analyses, there was little evidence of association of partner’s smoking with HDP, both overall and separately for gestational hypertension and preeclampsia (0.97, 0.81 to 1.14, P=0.682 for HDP) (Figure 2, supplementary tables 8-10).

### Beverage type analysis

Beer drinkers were much more likely to smoke before and during pregnancy and less likely to be married than non-drinkers (supplementary table 11). Those who drank wine during pregnancy were older, more likely to be white and much more likely to have a degree than non-drinkers (supplementary table 12). We compared risk of HDP stratified by beverage type (Figure 3). Point estimates were consistently more extreme for wine compared to beer, and the former but not the latter showed evidence of an association with lower HDP risk, although confidence intervals overlap between these analyses (supplementary tables 13 and 14).

**Figure 3.**
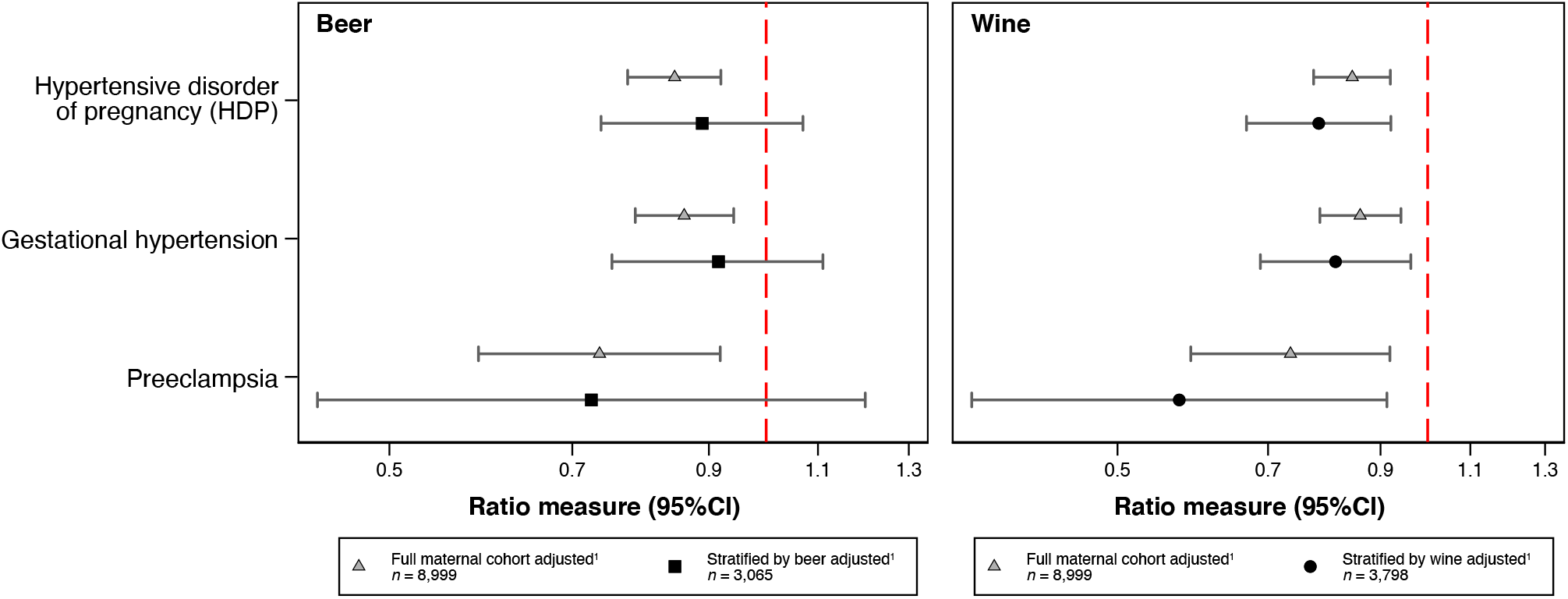
Beverage type analysis showing associations between beer and wine consumption and HDP, gestational hypertension and preeclampsia. Findings from the full maternal cohort, showing the ratio measure for one category increase of maternal drinking shown in both panels, adjusted for confounders. Below each finding from the full maternal cohort are the results of stratifying by beverage type showing the ratio measure for one category increase in beer or wine intake during pregnancy. ^1^ Adjusted for age, BMI, pre- and during pregnancy smoking (binary), parity, ethnicity, educational attainment, and marital status.

To understand drinking patterns in beer and wine drinkers during pregnancy, we compared binge drinking and reported use of other alcoholic drinks (spirits/other). Beer drinkers were more likely to report binge drinking during pregnancy and although there were no differences in intake of other drinks between beer and wine drinkers, there was significantly more missing data for these questions for beer than wine drinkers (supplementary table 15). Differing distributions of spirit intake and missing data between beer and wine demonstrates the difference in social patterning of wine and beer drinking.

### Full maternal cohort sensitivity analyses

Figure 4 summarises the findings from the primary analysis in the full maternal cohort overlaid on each of the three sensitivity analysis panels for reference. The sensitivity analyses suggested that differential exposure misclassification (outcome influencing reporting of the exposure), residual confounding by smoking, and potential poorer health of non-drinkers prior to pregnancy had little to no effect on our overall estimates (supplementary tables 16-18).

**Figure 4.**
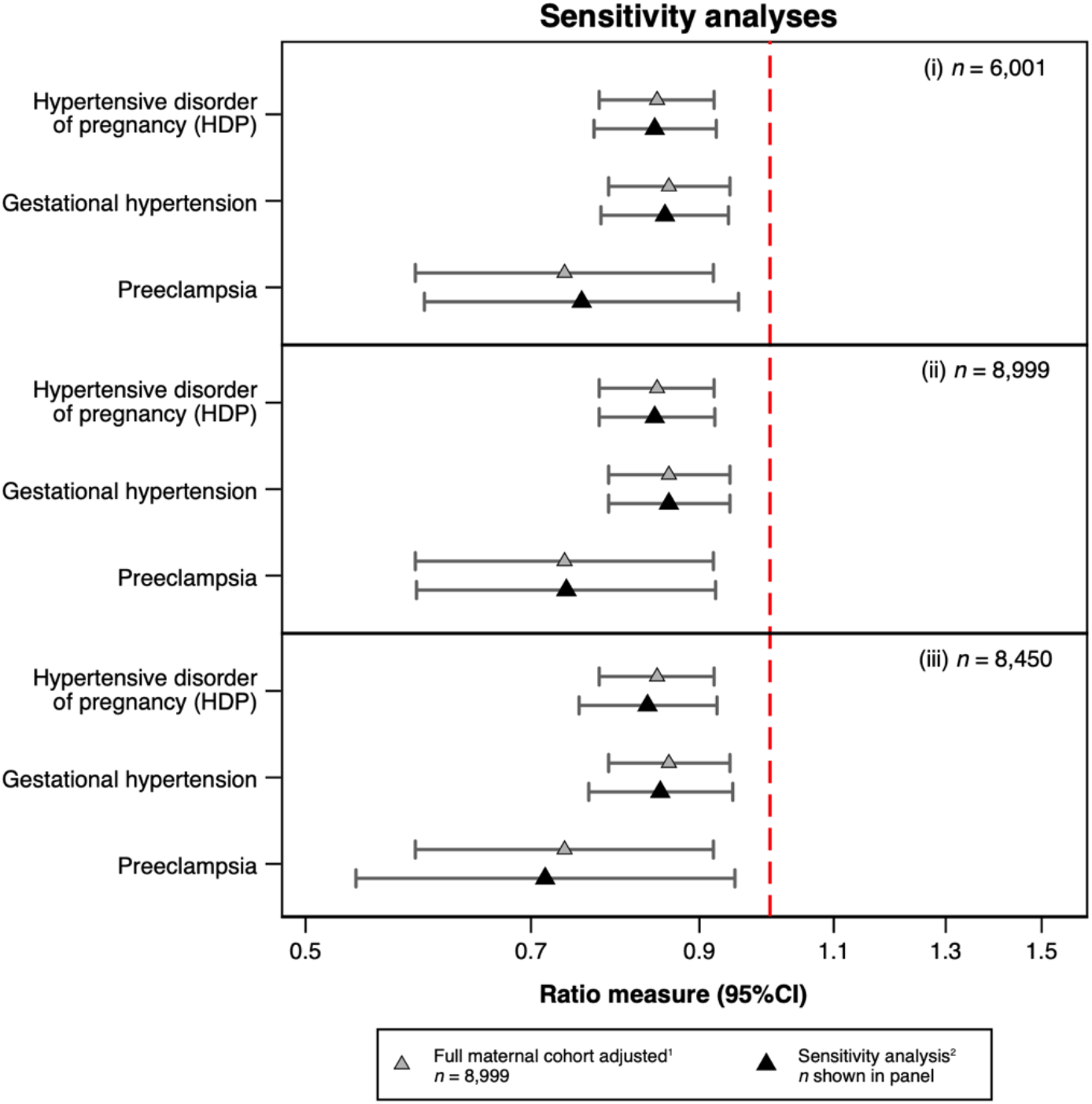
Sensitivity analyses showing associations between alcohol consumption during pregnancy and HDP, gestational hypertension, and preeclampsia. (i) Excluding those who reported their alcohol drinking after 20 weeks’ gestation (ii) Using number of cigarettes per day (0, 1–4, 5–9, 10–14, 15–19, 20–29 and 30+) (iii) Excluding those who reported abstaining from alcohol prior to their pregnancy ^1^ Adjusted for age, BMI, pre- and during pregnancy smoking (binary), parity, ethnicity, educational attainment, and marital status. ^2^ Adjusted for age, BMI, pre- and during pregnancy smoking (binary in model (i) and (iii), categorical in model (ii)), parity, ethnicity, educational attainment, and marital status The denominator in each analysis is different depending on the criteria of the sensitivity analysis; for example,(i) was performed in those participants from the full maternal cohort who had responded to questionnaire B prior to 20 weeks’ gestation (*n*=6,001).

## Discussion

We found that maternal alcohol intake during pregnancy was negatively associated with any HDP, both gestational hypertension and preeclampsia, which was also confirmed in multiple sensitivity analyses. In the negative control analysis, partner’s drinking was also inversely associated with maternal HDP, even after adjusting for maternal alcohol intake during pregnancy.

A (not yet peer-reviewed) recent systematic review identified an inverse association between alcohol intake during pregnancy and preeclampsia when stratifying by prospective studies, but not when including all eligible studies (13). In this review, only two of the included prospective studies used multivariable analyses to account for confounding. The first was a multi-country cohort study comparing those who quit drinking alcohol prior to 15 weeks’ gestation with those who did not drink alcohol found that this pattern of alcohol intake during pregnancy was associated with a decreased risk of preeclampsia (10). In the other, Iwama *et al*. observed HDP point estimates below one for those drinking almost no alcohol and less than 19 units of alcohol per week compared to none when adjusting for covariates, but with large standard errors and wide confidence intervals due to small numbers in the drinking groups (24). The largest included retrospective study was a US record linkage analysis which found that one to two drinks per week prenatally were negatively associated with preeclampsia compared to none in minimally adjusted models (adjusted OR 0.82, 95% CI 0.74 to 0.90) (7). Our findings were consistent with the results of these studies examining similar levels of alcohol intake. However, our unique take of running in parallel an analysis of partner’s exposure revealed that a causal effect is highly unlikely.

The main strength of the present study is that we uniquely applied a negative control design using partner’s alcohol intake during pregnancy. This provided a clearer insight into whether the association that was observed in the analysis of maternal alcohol intake was potentially causal, eventually concluding that shared confounding was a much more likely explanation. We additionally used smoking during pregnancy to validate this approach in the context of our data, and showed that the association between partner smoking and HDP attenuated considerably when adjusting for maternal smoking. This provided further support to our interpretation that shared (residual) confounding may be driving our inverse estimates of the prenatal alcohol-HDP association.

Confounding by SEP poses an additional risk to inferring causality for the prenatal alcohol-HDP association results. The J-shaped curve is well-discussed in alcohol and cardiovascular health epidemiology, where low-to-moderate amounts of alcohol intake appear to confer cardioprotective effects (25). Whether this is causal, or a result of confounding by SEP is hotly debated. A large Mendelian randomisation meta-analysis, which is less prone to the limitations suffered by traditional observational analyses, found that those with alleles associated with lower alcohol intake had a more favourable cardiovascular profile than those without the variant, suggesting that the J-shaped curve may be a result of confounding by SEP (5). Given that types of beverages consumed are also socially patterned, granular data on beer and wine intake in our cohort allowed us to run additional analyses separately for participants who drank beer and not wine (and vice versa). Investigating beer and wine separately in a beverage type analysis can be seen as an alternative method to capture some residual socioeconomic confounding that may not have been adequately accounted for by highest maternal educational attainment. The beverage type analysis showed wine to have a stronger inverse association with HDP than beer, which is consistent with the often-reported protective effect observed for wine drinking and health outcomes (26). The most likely explanation for wine’s protective effect on health is that wine drinkers share other characteristics that convey this benefit over non-wine drinkers, inadequately accounted for in our beverage-type analysis and previously published studies, as opposed to a causal effect.

We were able to run a number of sensitivity analyses to address the possibility of different types of bias explaining our results. First, we excluded those who reported abstaining from alcohol prior to their pregnancy due to potential differences in risk of the outcome between non-drinkers and drinkers prior to pregnancy (27, 28) to reduce the impact of reverse causation (ill-health causing drinking behaviour, i.e., abstaining from drinking). Given the potential for recall bias thus differential exposure misclassification, we restricted the cohort to women who had reported their drinking habits prior to 20 weeks’ gestation (the earliest point in pregnancy that HDP can be diagnosed). The findings from both of these sensitivity analyses mirrored the primary analysis and suggested that behaviour modification based on health and behaviour reporting based on pregnancy progression weren’t playing a significant role in the observed association from the primary analysis. However, it remains important to consider the potential effect that discussions with healthcare professionals during early antenatal appointment could have on behaviour or reporting of alcohol use. Smoking has been repeatedly shown to be associated with decreased HDP risk (12) and is correlated with alcohol use, so residual confounding by smoking behaviour could introduce bias, strengthening the inverse association. Using multiple measures of smoking throughout pregnancy from multiple questionnaires, we were able to mitigate as much of the confounding by smoking as permitted by the data we have in ALSPAC on prenatal smoking.

In addition to our negative control exposure analysis and multiple sensitivity analyses, a notable strength is the prospective collection of alcohol intake which wards against recall bias. The collection of outcome data on HDP from obstetric records improved reliability and reduced amounts of missing data. This study did have some limitations. Despite the large sample size, the number of women with preeclampsia was modest, though in line with other published estimates (29), supporting generalisability of this study. Exposure misclassification may have been an issue in this study, especially if heavy drinkers under-reported their alcohol use due to desirability bias. Although we used baseline variables in ALSPAC, thus participant attrition was relatively low, complete cases included in the analysis were less likely to drink or smoke during pregnancy, more likely to be older, married, and have higher educational attainment affecting internal validity.

In conclusion, we found that both maternal and partner’s alcohol intake during pregnancy were inversely associated with risk of any HDP, including gestational hypertension and preeclampsia. Our negative control analysis and the stronger protective effect of wine (as opposed to beer) compared to not drinking during pregnancy suggests that the association is not likely to reflect a direct, causal effect of maternal alcohol intake.

### Perspectives

Women who drink alcohol during pregnancy appear to be at a lower risk of developing HDP, however it has been deemed unclear as to whether this association is causal or due to artefacts (confounding, other biases). Three large cohort studies have reported conflicting findings, with smaller studies consistently reporting lower alcohol intake in preeclamptic pregnancies compared to healthy pregnancies. The present study found an association between alcohol intake in pregnancy and decreased risk of HDP, overall and stratified by gestational hypertension and preeclampsia. Using multivariable modelling and a negative control design, we interpret this negative association as the result of residual confounding, implicating SEP as an important factor, rather than a causal effect. These findings should be triangulated with those obtained using different methods and analytical strategies, e.g. Mendelian randomisation, to provide clarity on the true nature of this association. It is important that these findings are considered in the debate surrounding alcohol intake in pregnancy, alongside the other known harms to both mother and fetus (30).

### Novelty and significance

#### What is new?

- A negative association between alcohol intake during pregnancy and hypertensive disorders of pregnancy (HDP) has been previously suggested in a limited number of observational studies at high risk of confounding bias. We have attempted to answer this question using a robust causal inference approach.
- It is unclear whether the decreased risk of maternal HDP associated with drinking alcohol during pregnancy is caused by alcohol itself, or other factors correlated with drinking alcohol such as smoking.

### What is relevant?

- We found that maternal alcohol intake during pregnancy was associated with a decreased risk of developing HDP, as was partner drinking, even after mutually adjusting for confounders and maternal drinking.

### Summary

We conclude that the negative association observed between drinking alcohol during pregnancy and risk of HDP is a result of common environmental exposures shared between pregnant women and their partner’s, as opposed to a causal effect, and this warrants further investigation including using different study designs.

## Data Availability

Source code can be found here https://github.com/flozoemartin/MP2, ALSPAC data not publicly available.

## Funding

This research was performed in the UK Medical Research Council Integrative Epidemiology Unit (grant number: MC_UU_00011/7) and also supported by the National Institute for Health Research (NIHR) Bristol Biomedical Research Centre at University Hospitals Bristol National Health Service (NHS) Trust and the University of Bristol. The Wellcome Trust also funds FZM’s PhD studentship (grant reference: 218495/Z/19/Z) and LZ was supported by a UK MRC fellowship (grant number: G0902144). AF was supported by an MRC personal fellowship (grant reference: MR/M009351/1). The UK Medical Research Council and the Wellcome Trust (grant reference: 217065/Z/19/Z) and the University of Bristol provide core support for ALSPAC. Further details of grant funding for ALSPAC are available on their website.

## Acknowledgements

We are extremely grateful to all the families who took part in this study, the midwives for their help in recruiting them and the whole ALSPAC team, which includes interviewers, computer and laboratory technicians, clerical workers, research scientists, volunteers, managers, receptionists, and nurses.

## Notes

### Competing Interest Statement

The authors have declared no competing interest.

### Author Declarations

ALSPAC Ethics and Law Committee and local Research Ethics Committee (North Somerset and South Bristol)

