## Supplementary tables for "Alcohol intake and hypertensive disorders of pregnancy: a negative control analysis in the ALSPAC cohort"

### SUPPLEMENTARY MATERIAL

Supplementary table 1. Summary of the variables in the full cohort used in the full analysis (*n*=15,442), those in the complete cohort (*n*=8,999) and excluded, incomplete cases for exposure, outcomes, and covariates.

| Characteristic | Categories | Available data (n = 15,442) *n* (%) | Categorical data (n = 15,442) *n* (%) | Complete records (n = 8,999) *n* (%) | Records with incomplete data *n* | Excluded *n* (%) |
|---|---|---|---|---|---|---|
| Exposure: Alcohol use in pregnancy | Non-drinker | 12,353 (80) | 3,195 (26) | 2,415 (27) | 3,022 | 680 (23) |
| | Low-to-moderate | | 6,439 (52) | 4,696 (52) | | 1,574 (52) |
| | Heavy | | 2,719 (22) | 1,888 (21) | | 768 (25) |
| Age | Under 25 | 13,877 (90) | 3,337 (24) | 1,719 (19) | 4,546 | 1,567 (34) |
| | 25 and over | | 10,540 (76) | 7,280 (81) | | 2,979 (66) |
| Body mass index (BMI) | Underweight | 11,504 (75) | 577 (5) | 425 (5) | 2,173 | 143 (7) |
| | Normal | | 8,562 (74) | 6,767 (75) | | 1,581 (73) |
| | Overweight | | 1,734 (15) | 1,355 (15) | | 318 (15) |
| | Obese | | 631 (6) | 452 (5) | | 131 (6) |
| Smoking before pregnancy | Non-smoker | 13,173 (85) | 8,724 (66) | 6,121 (68) | 3,842 | 2,351 (61) |
| | Smoker | | 4,449 (34) | 2,878 (32) | | 1,491 (39) |
| Smoking during pregnancy | Non-smoker | 11,994 (78) | 8,309 (69) | 6,703 (75) | 2,663 | 1,335 (50) |
| | Smoker | | 3,685 (31) | 2,296 (25) | | 1,328 (50) |
| Parity | Nulliparous | 12,940 (84) | 5,805 (45) | 4,006 (45) | 3,609 | 1,619 (45) |
| | Multiparous | | 7,135 (55) | 4,993 (55) | | 1,990 (55) |
| Ethnicity | White | 12,231 (79) | 11,910 (97) | 8,834 (98) | 2,900 | 2,747 (95) |
| | Non-white | | 321 (3) | 165 (2) | | 153 (5) |
| Educational attainment | A levels or less | 12,322 (80) | 10,736 (87) | 7,754 (86) | 2,991 | 2,699 (90) |
| | Degree | | 1,586 (13) | 1,245 (14) | | 292 (10) |
| Marital status | Not currently married | 13,528 (88) | 3,523 (26) | 2,000 (22) | 4,197 | 1,448 (35) |
| | Married | | 10,005 (74) | 6,999 (78) | | 2,749 (65) |
| HDP | Normotensive | 13,682 (89) | 11,448 (84) | 7,509 (83) | 4,351 | 3,605 (83) |
| | Gestational hypertension | | 1,937 (14) | 1,308 (15) | | 631 (15) |
| | Preeclampsia | | 297 (2) | 182 (2) | | 115 (3) |

Supplementary table 2. Predictors of being a complete case in available data for each variable

| Characteristic | Category | Crude OR (95%CI) |
|---|---|---|
| Exposure: Alcohol use in pregnancy | Non-drinker | 1.00 (referent) |
| | Low-to-moderate | 0.84 (0.76 to 0.93) |
| | Heavy | 0.70 (0.62 to 0.78) |
| Age | Under 25 | 1.00 (referent) |
| | 25 and over | 2.24 (2.07 to 2.43) |
| Body mass index (BMI) | Underweight | 1.00 (referent) |
| | Normal | 1.44 (1.18 to 1.75) |
| | Overweight | 1.46 (1.15 to 1.81) |
| | Obese | 1.23 (0.94 to 1.62) |
| Smoking before pregnancy | Non-smoker | 1.00 (referent) |
| | Smoker | 0.73 (0.68 to 0.79) |
| Smoking during pregnancy | Non-smoker | 1.00 (referent) |
| | Smoker | 0.34 (0.31 to 0.37) |
| Parity | Nulliparous | 1.00 (referent) |
| | Multiparous | 1.00 (0.93 to 1.08) |
| Ethnicity | White | 1.00 (referent) |
| | Non-white | 0.33 (0.27 to 0.41) |
| Educational attainment | A-levels or lower | 1.00 (referent) |
| | University degree | 1.48 (1.29 to 1.69) |
| Marital status | Not currently married | 1.00 (referent) |
| | Married | 1.84 (1.70 to 1.99) |
| Outcome: HDP | Normotensive | 1.00 (referent) |
| | HDP | 0.92 (0.83 to 1.01) |

### Supplementary table 3. Maternal alcohol consumption during pregnancy and HDP in full cohort (*n*=8,999)

| Maternal outcome | Maternal alcohol use during pregnancy | | | Unadjusted model | | Adjusted model | |
|---|---|---|---|---|---|---|---|
| | Heavy *n* (%) | Low-to-moderate *n* (%) | None *n* (%) | OR[a] (95%CI) | *p*-value | OR[a,b] (95%CI) | *p*-value |
| **HDP** – *n* complete cases | **1,888** | **4,696** | **2,415** | - | - | - | - |
| Yes | 259 (13.7) | 765 (16.3) | 466 (19.3) | 0.82 (0.75 to 0.88) | <0.001 | 0.85 (0.78 to 0.92) | <0.001 |
| | | | | RR[c] (95%CI) | *p*-value | RR[b,c] (95%CI) | *p*-value |
| **Gestational hypertension** – *n* complete cases | **1,888** | **4,696** | **2,415** | - | - | - | - |
| Yes | 229 (12.1) | 681 (14.5) | 398 (16.5) | 0.83 (0.76 to 0.91) | <0.001 | 0.86 (0.79 to 0.94) | 0.001 |
| **Preeclampsia** – *n* complete cases | **1,888** | **4,696** | **2,415** | - | - | - | - |
| Yes | 30 (1.6) | 84 (1.8) | 68 (2.8) | 0.70 (0.56 to 0.87) | 0.001 | 0.74 (0.59 to 0.92) | 0.007 |

[a] Odds ratio generated using logistic regression
[b] Adjusted for covariates: maternal age at delivery, maternal BMI (12 weeks' gestation), pre-pregnancy smoking (binary), smoking during pregnancy (binary), parity (0, 1, 2 or ≥3), maternal ethnicity (white or non-white), maternal education (32 weeks' gestation) and marital status
[c] Relative risk ratio generated by multinomial logistic regression

### Supplementary table 4. Maternal alcohol consumption during pregnancy and HDP in negative control cohort (*n*=5,376)

| Maternal outcome | Maternal alcohol use during pregnancy | | | Unadjusted model | | Adjusted model | | Mutually adjusted model | |
|---|---|---|---|---|---|---|---|---|---|
| | Heavy *n* (%) | Low-to-moderate *n* (%) | None *n* (%) | OR[a] (95%CI) | *p*-value | OR[a,b] (95%CI) | *p*-value | OR[a,b,d] (95%CI) | *p*-value |
| **HDP** – *n* complete cases | **1,030** | **2,872** | **1,474** | - | - | - | - | - | - |
| Yes | 153 (14.9) | 493 (17.2) | 310 (21.0) | 0.80 (0.72 to 0.89) | <0.001 | 0.84 (0.75 to 0.94) | 0.002 | 0.86 (0.77 to 0.96) | 0.008 |
| | | | | RR[c] (95%CI) | *p*-value | RR[b,c] (95%CI) | *p*-value | RR[b,c,d] (95%CI) | *p*-value |
| **Gestational hypertension** – *n* complete cases | **1,030** | **2,872** | **1,474** | - | - | - | - | - | - |
| Yes | 137 (13.3) | 434 (15.1) | 266 (18.1) | 0.82 (0.74 to 0.92) | <0.001 | 0.85 (0.76 to 0.96) | 0.007 | 0.87 (0.78 to 0.98) | 0.026 |
| **Preeclampsia** – *n* complete cases | **1,030** | **2,872** | **1,474** | - | - | - | - | - | - |
| Yes | 16 (1.6) | 59 (2.1) | 44 (3.0) | 0.69 (0.52 to 0.90) | 0.007 | 0.74 (0.55 to 0.98) | 0.033 | 0.75 (0.56 to 0.99) | 0.045 |

[a] Odds ratio generated using logistic regression
[b] Adjusted for covariates: maternal age at delivery, maternal BMI (12 weeks' gestation), pre-pregnancy smoking (binary), smoking during pregnancy (binary), parity (0, 1, 2 or ≥3), maternal ethnicity (white or non-white), maternal education (32 weeks' gestation) and marital status
[c] Relative risk ratio generated by multinomial logistic regression
[d] Mutually adjusted for covariates plus partner's alcohol consumption during pregnancy

### Stratifying heavy drinking by binge drinking

As described in the methods section, the heavy drinking category was further stratified by binge and non-binge drinkers. It has been shown that different forms of drinking are more frequent in different socioeconomic groups: those who drink little and often (for example, one glass of wine every night) and those who drink heavily once a week, are both heavy drinkers in our analysis. However, they have been shown in this sample (supplementary table 5) to be characteristically different from one another; we deduced that this may have impacted on their underlying risk of outcome. There was a slight attenuation of the protective effect for heavy binge drinkers vs heavy non-binge drinkers when comparing with non-drinkers (as per the primary anlaysis), but the direction of effect was the same for all three outcomes and the intervals did not include one (supplementary table 6).

**Supplementary table 5. Characteristics of participants by categories of alcohol consumption during pregnancy, heavy stratified by binge and non-binge**

| | None | Low-to-moderate | Heavy non-binge | Heavy binge |
|---|---|---|---|---|
| | **2,415** | **4,696** | **348** | **1,540** |
| **Age at delivery** | | | | |
| Mean, years (SD[a]) | 27.7 (4.7) | 28.9 (4.6) | 30.4 (4.7) | 28.4 (4.9) |
| **BMI[b] (12 weeks' gestation)** | | | | |
| Mean, kg/m$^2$ (SD[a]) | 23.0 (4.0) | 22.7 (3.6) | 22.5 (3.3) | 23.2 (3.0) |
| **Smoking** | | | | |
| Any pre-pregnancy, *n* (%) | 705 (29.2) | 1,299 (27.7) | 117 (33.6) | 757 (49.2) |
| Any during pregnancy, *n* (%) | 552 (22.9) | 991 (21.1) | 91 (26.2) | 662 (43.0) |
| **Parity (18 weeks' gestation)** | | | | |
| Multiparous, *n* (%) | 1,280 (53.0) | 2,589 (55.1) | 199 (57.2) | 925 (60.1) |
| **Ethnicity (32 weeks' gestation)** | | | | |
| Non-white, *n* (%) | 61 (2.5) | 80 (1.7) | 4 (1.2) | 20 (1.3) |
| **Educational attainment (32 weeks' gestation)** | | | | |
| University degree, *n* (%) | 232 (9.6) | 792 (16.9) | 102 (29.3) | 119 (7.7) |
| **Marital status** | | | | |
| Married, *n* (%) | 1,904 (78.8) | 3,792 (80.8) | 273 (78.5) | 1,030 (66.9) |

[a] Standard deviation
[b] Body mass index

Supplementary table 6. Maternal alcohol consumption during pregnancy and HDP, expanding heavy drinking to binge and non-binge (*n*=8,999), including alcohol exposure as a categorical exposure

| Type of drinking | Maternal outcome | Unadjusted OR[a] (95%CI) | *p*-value | Adjusted OR[a,b] (95%CI) | *p*-value |
|---|---|---|---|---|---|
| | HDP *n* (%) | - | - | - | - |
| None | 466/2,415 (19.3) | 1.00 (reference) | - | 1.00 (reference) | - |
| Low-to-moderate | 765/4,696 (16.3) | 0.81 (0.72 to 0.92) | 0.002 | 0.85 (0.74 to 0.97) | 0.019 |
| Heavy non-binge | 43/348 (12.4) | 0.59 (0.42 to 0.82) | 0.002 | 0.64 (0.45 to 0.90) | 0.012 |
| Heavy binge | 216/1,540 (14.0) | 0.68 (0.57 to 0.81) | <0.001 | 0.73 (0.60 to 0.88) | 0.001 |
| | | Unadjusted RR[c] (95%CI) | *p*-value | Adjusted RR[b,c] (95%CI) | *p*-value |
| | Gestational hypertension | | | | |
| None | 398/2,415 (16.5) | 1.00 (reference) | - | 1.00 (reference) | - |
| Low-to-moderate | 681/4,696 (14.5) | 0.85 (0.74 to 0.97) | 0.017 | 0.88 (0.77 to 1.02) | 0.083 |
| Heavy non-binge | 36/348 (10.3) | 0.58 (0.40 to 0.83) | 0.003 | 0.62 (0.43 to 0.90) | 0.013 |
| Heavy binge | 193/1,540 (12.5) | 0.71 (0.59 to 0.86) | <0.001 | 0.76 (0.63 to 0.92) | 0.006 |
| | Preeclampsia | - | - | - | - |
| None | 68/2,415 (2.8) | 1.00 (reference) | - | 1.00 (reference) | - |
| Low-to-moderate | 84/4,696 (1.8) | 0.61 (0.44 to 0.85) | 0.003 | 0.66 (0.48 to 0.93) | 0.017 |
| Heavy non-binge | 7/348 (2.0) | 0.66 (0.30 to 1.45) | 0.297 | 0.74 (0.33 to 1.66) | 0.463 |
| Heavy binge | 23/1,540 (1.5) | 0.50 (0.31 to 0.80) | 0.004 | 0.54 (0.33 to 0.87) | 0.013 |

[a] Odds ratio generated using logistic regression
[b] Adjusted for covariates: maternal age at delivery, maternal BMI (12 weeks' gestation), pre-pregnancy smoking (binary), smoking during pregnancy (binary), parity (0, 1, 2 or ≥3), maternal ethnicity (white or non-white), maternal education (32 weeks' gestation) and marital status
[c] Relative risk ratio generated by multinomial logistic regression

Supplementary table 7. Partner's alcohol consumption during pregnancy and maternal HDP in negative control cohort (*n*=5,376)

| Maternal outcome | Partner's alcohol use during pregnancy | | | Unadjusted model | | Adjusted model | | Mutually adjusted model | |
|---|---|---|---|---|---|---|---|---|---|
| | Heavy *n* (%) | Low-to-moderate *n* (%) | None *n* (%) | OR[a] (95%CI) | *p*-value | OR[a,b] (95%CI) | *p*-value | OR[a,b,d] (95%CI) | *p*-value |
| **HDP** – *n* complete cases | 1,292 | 3,943 | 141 | - | - | - | - | - | - |
| Yes | 196 (15.2) | 728 (18.5) | 32 (22.7) | 0.79 (0.68 to 0.92) | 0.002 | 0.79 (0.67 to 0.92) | 0.003 | 0.82 (0.70 to 0.97) | 0.018 |
| | | | | RR[c] (95%CI) | *p*-value | RR[b,c] (95%CI) | *p*-value | RR[b,c,d] (95%CI) | *p*-value |
| **Gestational hypertension** – *n* complete cases | 1,292 | 3,943 | 141 | - | - | - | - | - | - |
| Yes | 168 (13.0) | 642 (16.3) | 27 (19.2) | 0.78 (0.66 to 0.91) | 0.002 | 0.78 (0.66 to 0.92) | 0.003 | 0.81 (0.68 to 0.96) | 0.014 |
| **Preeclampsia** – *n* complete cases | 1,292 | 3,943 | 141 | - | - | - | - | - | - |
| Yes | 28 (2.2) | 86 (2.2) | 5 (3.6) | 0.87 (0.59 to 1.29) | 0.494 | 0.86 (0.58 to 1.30) | 0.479 | 0.95 (0.63 to 1.43) | 0.794 |

[a] Odds ratio generated using logistic regression
[b] Adjusted for covariates: partner's age at delivery, partner's BMI (12 weeks' gestation), pre-pregnancy smoking (binary), smoking during pregnancy (binary), parity (0, 1, 2 or ≥3), partner's ethnicity (white or non-white), partner's education (32 weeks' gestation) and marital status
[c] Relative risk ratio generated by multinomial logistic regression
[d] Mutually adjusted for covariates plus maternal alcohol consumption during pregnancy

### Supplementary table 8. Maternal smoking during pregnancy and HDP in full cohort (*n*=8,999)

| Maternal outcome | Maternal smoking during pregnancy | | Unadjusted model | | Adjusted model | |
|---|---|---|---|---|---|---|
| | Any *n* (%) | None *n* (%) | OR[a] (95%CI) | *p*-value | OR[a,b] (95%CI) | *p*-value |
| **HDP** – *n* complete cases | **2,296** | **6,703** | - | - | - | - |
| Yes | 301 (13.1) | 1,189 (17.7) | 0.70 (0.61 to 0.80) | <0.001 | 0.67 (0.53 to 0.86) | 0.001 |
| | | | RR[c] (95%CI) | *p*-value | RR[b,c] (95%CI) | *p*-value |
| **Gestational hypertension** – *n* complete cases | **2,296** | **6,703** | - | - | - | - |
| Yes | 271 (11.8) | 1,037 (15.5) | 0.72 (0.63 to 0.83) | <0.001 | 0.72 (0.56 to 0.93) | 0.011 |
| **Preeclampsia** – *n* complete cases | **2,296** | **6,703** | - | - | - | - |
| Yes | 30 (1.3) | 152 (2.3) | 0.55 (0.37 to 0.81) | 0.003 | 0.41 (0.23 to 0.74) | 0.003 |

[a] Odds ratio generated using logistic regression
[b] Adjusted for covariates: maternal age at delivery, maternal BMI (12 weeks' gestation), pre-pregnancy smoking (binary), smoking during pregnancy (binary), parity (0, 1, 2 or ≥3), maternal ethnicity (white or non-white), maternal education (32 weeks' gestation) and marital status
[c] Relative risk ratio generated by multinomial logistic regression

### Supplementary table 9. Maternal smoking during pregnancy and HDP in negative control cohort (*n*=5,376)

| Maternal outcome | Maternal smoking during pregnancy | | Unadjusted model | | Adjusted model | | Mutually adjusted model | |
|---|---|---|---|---|---|---|---|---|
| | Any *n* (%) | None *n* (%) | OR[a] (95%CI) | *p*-value | OR[a,b] (95%CI) | *p*-value | OR[a,b,d] (95%CI) | *p*-value |
| **HDP** – *n* complete cases | 1,169 | 4,207 | - | - | - | - | - | - |
| Yes | 159 (13.6) | 797 (18.9) | 0.67 (0.56 to 0.81) | <0.001 | 0.64 (0.52 to 0.79) | <0.001 | 0.66 (0.53 to 0.81) | <0.001 |
| | | | RR[c] (95%CI) | *p*-value | RR[b,c] (95%CI) | *p*-value | RR[b,c,d] (95%CI) | *p*-value |
| **Gestational hypertension** – *n* complete cases | 1,169 | 4,207 | - | - | - | - | - | - |
| Yes | 147 (12.6) | 690 (16.4) | 0.72 (0.59 to 0.87) | 0.001 | 0.70 (0.57 to 0.86) | 0.001 | 0.71 (0.57 to 0.89) | 0.003 |
| **Preeclampsia** – *n* complete cases | 1,169 | 4,207 | - | - | - | - | - | - |
| Yes | 12 (1.0) | 107 (2.5) | 0.38 (0.21 to 0.69) | 0.002 | 0.30 (0.16 to 0.56) | <0.001 | 0.32 (0.17 to 0.61) | 0.001 |

[a] Odds ratio generated using logistic regression
[b] Adjusted for covariates: maternal age at delivery, maternal BMI (12 weeks' gestation), pre-pregnancy smoking (binary), alcohol use during pregnancy (none, low-to-moderate or heavy), parity (0, 1, 2 or ≥3), maternal ethnicity (white or non-white), maternal education (32 weeks' gestation) and marital status
[c] Relative risk ratio generated by multinomial logistic regression
[d] Mutually adjusted for covariates plus partner's smoking during pregnancy

Supplementary table 10. Partner's smoking during pregnancy and maternal HDP in negative control cohort (*n*=5,376)

| Maternal outcome | Partner's smoking during pregnancy | | Unadjusted model | | Adjusted model | | Mutually adjusted model | |
|---|---|---|---|---|---|---|---|---|
| | Any *n* (%) | None *n* (%) | OR[a] (95%CI) | *p*-value | OR[a,b] (95%CI) | *p*-value | OR[a,b,d] (95%CI) | *p*-value |
| **HDP** – *n* complete cases | 1,815 | 3,561 | - | - | - | - | - | - |
| Yes | 298 (16.4) | 658 (18.5) | 0.87 (0.75 to 1.01) | 0.062 | 0.86 (0.73 to 1.00) | 0.056 | 0.97 (0.81 to 1.14) | 0.682 |
| | | | RR[c] (95%CI) | *p*-value | RR[b,c] (95%CI) | *p*-value | RR[b,c,d] (95%CI) | *p*-value |
| **Gestational hypertension** – *n* complete cases | 1,815 | 3,561 | - | - | - | - | - | - |
| Yes | 264 (14.6) | 573 (16.1) | 0.88 (0.75 to 1.03) | 0.119 | 0.884 (0.75 to 1.05) | 0.152 | 0.98 (0.82 to 1.17) | 0.779 |
| **Preeclampsia** – *n* complete cases | 1,815 | 3,561 | - | - | - | - | - | - |
| Yes | 34 (1.9) | 85 (2.4) | 0.77 (0.51 to 1.14) | 0.193 | 0.67 (0.44 to 1.03) | 0.067 | 0.89 (0.57 to 1.38) | 0.606 |

[a] Odds ratio generated using logistic regression
[b] Adjusted for covariates: partner's age at delivery, partner's BMI (12 weeks' gestation), pre-pregnancy smoking (binary), partner's alcohol use during pregnancy (none, low-to-moderate or heavy), parity (0, 1, 2 or ≥3), partner's ethnicity (white or non-white), partner's education (32 weeks' gestation) and marital status
[c] Relative risk ratio generated by multinomial logistic regression
[d] Mutually adjusted for covariates plus maternal smoking during pregnancy

Supplementary table 11. Characteristics of participants stratified by categories of beer consumption, excluding those who report any wine consumption.

| | No alcohol in pregnancy | Low-to-moderate beer consumption in pregnancy | Heavy beer consumption in pregnancy |
|---|---|---|---|
| | **2,415** | **458** | **192** |
| **Age at delivery** | | | |
| Mean, years (SD[a]) | 27.7 (4.68) | 27.6 (4.70) | 28.6 (5.1) |
| **BMI[b] (12 weeks' gestation)** | | | |
| Mean, kg/m$^2$ (SD[a]) | 23.0 (4.02) | 23.0 (4.0) | 22.8 (3.1) |
| **Smoking** | | | |
| Any pre-pregnancy, *n* (%) | 705 (29.2) | 220 (48.0) | 111 (57.8) |
| Any during pregnancy, *n* (%) | 552 (22.9) | 191 (41.7) | 102 (53.1) |
| **Parity (18 weeks' gestation)** | | | |
| Multiparous, *n* (%) | 1,280 (53.0) | 240 (52.4) | 128 (66.7) |
| **Ethnicity (32 weeks' gestation)** | | | |
| Non-white, *n* (%) | 61 (2.5) | 3 (0.7) | 6 (3.1) |
| **Educational attainment (32 weeks' gestation)** | | | |
| University degree, *n* (%) | 232 (9.6) | 38 (8.3) | 16 (8.3) |
| **Marital status** | | | |
| Married, *n* (%) | 1,904 (78.8) | 317 (69.2) | 110 (57.3) |

[a] Standard deviation
[b] Body mass index

Supplementary table 12. Characteristics of participants stratified by categories of wine consumption, excluding those who report any beer consumption.

| | No alcohol in pregnancy | Low-to-moderate wine consumption in pregnancy | Heavy wine consumption in pregnancy |
|---|---|---|---|
| | **2,415** | **1,152** | **231** |
| **Age at delivery** | | | |
| Mean, years (SD[a]) | 27.7 (4.68) | 29.7 (4.3) | 30.9 (4.6) |
| **BMI[b] (12 weeks' gestation)** | | | |
| Mean, kg/m$^2$ (SD[a]) | 23.0 (4.02) | 22.8 (3.4) | 22.6 (3.2) |
| **Smoking** | | | |
| Any pre-pregnancy, *n* (%) | 705 (29.2) | 300 (26.0) | 81 (35.1) |
| Any during pregnancy, *n* (%) | 552 (22.9) | 215 (18.7) | 62 (26.8) |
| **Parity (18 weeks' gestation)** | | | |
| Multiparous, *n* (%) | 1,280 (53.0) | 642 (55.7) | 147 (63.6) |
| **Ethnicity (32 weeks' gestation)** | | | |
| Non-white, *n* (%) | 61 (2.5) | 14 (1.2) | 1 (0.4) |
| **Educational attainment (32 weeks' gestation)** | | | |
| University degree, *n* (%) | 232 (9.6) | 235 (20.4) | 58 (25.1) |
| **Marital status** | | | |
| Married, *n* (%) | 1,904 (78.8) | 979 (85.0) | 192 (83.1) |

[a] Standard deviation
[b] Body mass index

Supplementary table 13. Maternal beer consumption during pregnancies and HDP restricted to those with beer and wine data available ($n$=3,065)

| Maternal outcome | Maternal beer drinking | | | Unadjusted model | | Adjusted model | |
|---|---|---|---|---|---|---|---|
| | Heavy *n* (%) | Low-to-moderate *n* (%) | None *n* (%) | OR (95%CI) | *p*-value | OR (95%CI) | *p*-value |
| **HDP** – *n* complete cases | **192** | **458** | **2,415** | - | - | - | - |
| Yes | 27 (14.1) | 72 (15.7) | 466 (19.3) | 0.81 (0.68 to 0.96) | 0.017 | 0.89 (0.74 to 1.07) | 0.217 |
| | | | | RR[c] (95%CI) | *p*-value | RR[b,c] (95%CI) | *p*-value |
| **Gestational hypertension** – *n* complete cases | **192** | **458** | **2,415** | - | - | - | - |
| Yes | 24 (12.5) | 64 (14.0) | 398 (16.5) | 0.83 (0.69 to 1.00) | 0.049 | 0.92 (0.75 to 1.11) | 0.378 |
| **Preeclampsia** – *n* complete cases | **192** | **458** | **2,415** | - | - | - | - |
| Yes | 3 (1.6) | 8 (1.8) | 68 (2.8) | 0.67 (0.41 to 1.08) | 0.102 | 0.73 (0.44 to 1.20) | 0.211 |

[a] Odds ratio generated using logistic regression
[b] Adjusted for covariates: maternal age at delivery, maternal BMI (12 weeks' gestation), pre-pregnancy smoking (binary), smoking during pregnancy (binary), parity (0, 1, 2 or ≥3), maternal ethnicity (white or non-white), maternal education (32 weeks' gestation) and marital status
[c] Relative risk ratio generated by multinomial logistic regression

### Supplementary table 14. Maternal wine consumption during pregnancies and HDP restricted to those with beer and wine data available (*n*=3,065)

| Maternal outcome | Maternal wine drinking | | | Unadjusted model | | Adjusted model | |
|---|---|---|---|---|---|---|---|
| | Heavy *n* (%) | Low-to-moderate *n* (%) | None *n* (%) | OR (95%CI) | *p*-value | OR (95%CI) | *p*-value |
| **HDP** – *n* complete cases | **231** | **1,152** | **2,415** | - | - | - | - |
| Yes | 29 (12.6) | 170 (14.8) | 466 (19.3) | 0.75 (0.64 to 0.87) | <0.001 | 0.78 (0.67 to 0.92) | 0.003 |
| | | | | RR[c] (95%CI) | *p*-value | RR[b,c] (95%CI) | *p*-value |
| **Gestational hypertension** – *n* complete cases | **231** | **1,152** | **2,415** | - | - | - | - |
| Yes | 27 (11.7) | 152 (13.2) | 398 (16.5) | 0.78 (0.67 to 0.91) | 0.002 | 0.81 (0.69 to 0.96) | 0.378 |
| **Preeclampsia** – *n* complete cases | **231** | **1,152** | **2,415** | - | - | - | - |
| Yes | 2 (0.9) | 18 (1.6) | 68 (2.8) | 0.53 (0.34 to 0.82) | 0.004 | 0.57 (0.36 to 0.91) | 0.019 |

[a] Odds ratio generated using logistic regression
[b] Adjusted for covariates: maternal age at delivery, maternal BMI (12 weeks' gestation), pre-pregnancy smoking (binary), smoking during pregnancy (binary), parity (0, 1, 2 or ≥3), maternal ethnicity (white or non-white), maternal education (32 weeks' gestation) and marital status
[c] Relative risk ratio generated by multinomial logistic regression

### Supplementary table 15. Binge drinking and consumption of additional forms of alcohol in beer (*n*=650) and wine (*n*=1,383) drinking groups.

| | Wine drinkers in pregnancy | Beer drinkers in pregnancy |
|---|---|---|
| | **1,383** | **650** |
| **Bingeing[a]** | | |
| Yes, *n* (%) | 380 (27.5) | 293 (45.1) |
| Missing, *n* (%) | 10 (0.7) | 7 (1.1) |
| **Other alcohol intake[b]** | | |
| Yes, *n* (%) | 67 (4.8) | 28 (4.3) |
| Missing, *n* (%) | 229 (16.6) | 258 (39.7) |

[a] Bingeing defined as drinking 4 or more drinks in one sitting
[b] Other alcohol intake defined as reporting intake of spirits or "other" alcohol during pregnancy

### Sensitivity analysis findings

#### Excluding respondent's post-20 weeks' gestation

Given the diagnosis of gestational hypertension or preeclampsia occurs at 20 weeks' gestation or later, we considered the potential for knowledge of the outcome to have influenced reporting of the exposure, had the questionnaire been filled out after a diagnosis could have been made. With this in mind, we restricted the analysis to those who had filled in the questionnaire prior to 20 weeks' gestation (thus excluding responses from the postpartum questionnaire regarding drinking habits in the last two months of pregnancy) (part (i) Figure 4). The protective effect persisted in both the logistic and multinomial models having restricted to participants responding prior to 20 weeks' gestation (adjusted relative risk ratio 0.85, 95% confidence interval 0.76 to 0.95, P-value=0.003 and 0.72, 0.54 to 0.95, P=0.020, for gestational hypertension and preeclampsia, respectively).

**Supplementary table 16. Maternal alcohol consumption during pregnancy and HDP excluding those who responded after 20 weeks' gestation (*n*=6,001) – (i) on Figure 4.**

| Maternal outcome | Maternal alcohol use during pregnancy | | | Unadjusted model | | Adjusted model | |
|---|---|---|---|---|---|---|---|
| | Heavy *n* (%) | Low-to-moderate *n* (%) | None *n* (%) | OR[a] (95%CI) | *p*-value | OR[a,b] (95%CI) | *p*-value |
| **HDP** – *n* complete cases | **1,123** | **2,887** | **1,991** | - | - | - | - |
| Yes | 156 (13.9) | 462 (16.0) | 393 (19.7) | 0.80 (0.73 to 0.89) | <0.001 | 0.83 (0.75 to 0.92) | 0.001 |
| | | | | RR[c] (95%CI) | *p*-value | RR[b,c] (95%CI) | *p*-value |
| **Gestational hypertension** – *n* complete cases | **1,123** | **2,887** | **1,991** | - | - | - | - |
| Yes | 142 (12.6) | 412 (14.3) | 343 (17.2) | 0.82 (0.74 to 0.91) | <0.001 | 0.85 (0.76 to 0.95) | 0.003 |
| **Preeclampsia** – *n* complete cases | **1,123** | **2,887** | **1,991** | - | - | - | - |
| Yes | 14 (1.3) | 50 (1.7) | 50 (2.5) | 0.68 (0.51 to 0.89) | 0.005 | 0.72 (0.54 to 0.95) | 0.020 |

[a] Odds ratio generated using logistic regression
[b] Adjusted for covariates: maternal age at delivery, maternal BMI (12 weeks' gestation), pre-pregnancy smoking (binary), smoking during pregnancy (binary), parity (0, 1, 2 or ≥3), maternal ethnicity (white or non-white), maternal education (32 weeks' gestation) and marital status
[c] Relative risk ratio generated by multinomial logistic regression

#### Stratifying smoking during pregnancy

It has been shown on multiple occasions that smoking during pregnancy is associated with a protective effect for preeclampsia (1, 2). We also observed this in ALSPAC, using smoking during pregnancy and risk of HDP to corroborate our findings for the partner's alcohol negative control analysis (Figure 2). Given that smoking is associated with drinking alcohol during pregnancy it was important to reduce any residual confounding not accounted for by using a binary smoking covariate. Having categorised smoking during pregnancy into average number per day (0, 1–4, 5–9, 10–14, 15–19, 20–29 and 30+), we observed no difference in effect from the model that adjusted for binary smoking during pregnancy (any or none) (adjusted odds ratio 0.85, 95% confidence interval 0.78 to 0.92, P-value<0.001) (part (ii), Figure 4).

**Supplementary table 17. Maternal alcohol consumption during pregnancy and HDP using a more specific categorical smoking variable (*n*=8,999) – (ii) on Figure 4.**

| Maternal outcome | Maternal alcohol use during pregnancy | | | Adjusted model (binary smoking during pregnancy) | | Adjusted model (categorical smoking during pregnancy) | |
|---|---|---|---|---|---|---|---|
| | **Heavy *n* (%)** | **Low-to-moderate *n* (%)** | **None *n* (%)** | **OR[a,b] (95%CI)** | ***p*-value** | **OR[a,b] (95%CI)** | ***p*-value** |
| **HDP** – *n* complete cases | **1,888** | **4,696** | **2,415** | - | - | - | - |
| Yes | 259 (13.7) | 765 (16.3) | 466 (19.3) | 0.85 (0.78 to 0.92) | <0.001 | 0.85 (0.78 to 0.92) | <0.001 |
| | | | | **RR[b,c] (95%CI)** | ***p*-value** | **RR[b,c] (95%CI)** | ***p*-value** |
| **Gestational hypertension** – *n* complete cases | **1,888** | **4,696** | **2,415** | - | - | - | - |
| Yes | 229 (12.1) | 681 (14.5) | 398 (16.5) | 0.86 (0.79 to 0.94) | 0.001 | 0.86 (0.79 to 0.94) | 0.001 |
| **Preeclampsia** – *n* complete cases | **1,888** | **4,696** | **2,415** | - | - | - | - |
| Yes | 30 (1.6) | 84 (1.8) | 68 (2.8) | 0.74 (0.59 to 0.92) | 0.007 | 0.74 (0.59 to 0.92) | 0.007 |

[a] Odds ratio generated using logistic regression
[b] Adjusted for covariates: maternal age at delivery, maternal BMI (12 weeks' gestation), pre-pregnancy smoking (binary), smoking during pregnancy, parity (0, 1, 2 or ≥3), maternal ethnicity (white or non-white), maternal education (32 weeks' gestation) and marital status
[c] Relative risk ratio generated by multinomial logistic regression

### Excluding abstainers prior to pregnancy

It has been shown that those who abstain from alcohol outside of pregnancy are characteristically different and have exhibited different risks of morbidity and mortality compared with their drinking counterparts (3, 4). With the exchangeability in question between this group and those who did not abstain before pregnancy and their risk of the outcome, we performed a sensitivity analysis by which we excluded those women who had reported to have abstained from alcohol prior to pregnancy. Other than slightly attenuating the effect, probably due to decreased power following their exclusion, both the crude and adjusted models concluded a reduction in relative risk, following alcohol use in pregnancy, of both gestational hypertension and preeclampsia (adjusted relative risk ratio 0.86, 95% confidence interval 0.78 to 0.95, P-value=0.001 and 0.76, 0.60 to 0.95, P=0.019, respectively) (part (iii), Figure 4).

**Supplementary table 18. Maternal alcohol consumption and HDP excluding abstainers prior to pregnancy (*n*=8,450) – (iii) on Figure 4.**

| Maternal outcome | Maternal alcohol use during pregnancy | | | Unadjusted model | | Adjusted model | |
|---|---|---|---|---|---|---|---|
| | Heavy *n* (%) | Low-to-moderate *n* (%) | None *n* (%) | OR[a] (95%CI) | *p*-value | OR[a,b] (95%CI) | *p*-value |
| **HDP** – *n* complete cases | **1,881** | **4,663** | **1,906** | - | - | - | - |
| Yes | 257 (13.7) | 763 (16.4) | 375 (19.7) | 0.80 (0.74 to 0.88) | <0.001 | 0.84 (0.77 to 0.92) | <0.001 |
| | | | | RR[c] (95%CI) | *p*-value | RR[b,c] (95%CI) | *p*-value |
| **Gestational hypertension** – *n* complete cases | **1,881** | **4,663** | **1,906** | - | - | - | - |
| Yes | 227 (12.1) | 679 (14.6) | 321 (16.8) | 0.82 (0.75 to 0.90) | <0.001 | 0.86 (0.78 to 0.94) | 0.001 |
| **Preeclampsia** – *n* complete cases | **1,881** | **4,663** | **1,906** | - | - | - | - |
| Yes | 30 (1.6) | 84 (1.8) | 54 (2.8) | 0.70 (0.56 to 0.89) | 0.003 | 0.76 (0.60 to 0.95) | 0.019 |

[a] Odds ratio generated using logistic regression
[b] Adjusted for covariates: maternal age at delivery, maternal BMI (12 weeks' gestation), pre-pregnancy smoking (binary), smoking during pregnancy (binary), parity (0, 1, 2 or ≥3), maternal ethnicity (white or non-white), maternal education (32 weeks' gestation) and marital status
[c] Relative risk ratio generated by multinomial logistic regression
